# INFLUENCE OF AVAILABILITY, BARRIERS TO ACCESSIBILITY, AND UTILIZATION OF MENTAL HEALTH SERVICES ON PSYCHOLOGICAL DISTRESS STATUS OF UNDERGRADUATE STUDENTS AT UNITED STATES INTERNATIONAL UNIVERSITY-AFRICA, KENYA

**DOI:** 10.1101/2023.05.05.23289570

**Authors:** Melvin A. Wao, Calvin A. Omolo, Eliab Some, Michael Kihara, Gladys Njoroge

## Abstract

**Background:** Psychological distress is prevalent among university students worldwide. Research shows that there are inadequate efforts being made to improve the mental health of university students and there is low level of accessibility of university students to mental health services. This study aimed to determine the relationship between availability of mental health services, barriers to access to the services, and utilization of the services, on psychological distress status of undergraduate students at United States International University-Africa, Kenya

**Methods:** The research was conducted using a mixed methods research approach. Specifically, an exploratory sequential mixed methods research design was employed, including a cross-sectional survey and key informant interviews. The sample population was 249 undergraduate students at USIU-A, collected using cluster and stratified sampling procedure. Kessler Psychological Distress Scale (K10) was used to determine psychological distress status.

**Results:** This study found 76.8% of undergraduate students suffer from psychological distress with highest prevalence among seniors. Majority of students were aware of available sources of mental health services with three most frequently cited sources by students including counselor (87%), social support (84%), and peer counselor (80%). It was found that psychological distress status varies by students’ awareness of availability of psychologists or personal coping strategist. Association between barriers of accessibility and utilization of mental health services to psychological distress status of students i.e., peer stigma, societal stigma, and self-sufficiency.

**Conclusion:** At USIU-A, majority of students suffer from psychological distress. Whereas several sources of mental health services are available at the institution, a good number prefer to seek informal mental health services and two major barriers to mental health service accessibility include attitudinal barriers and stigma.

**GRAPHICAL ABSTRACT:** 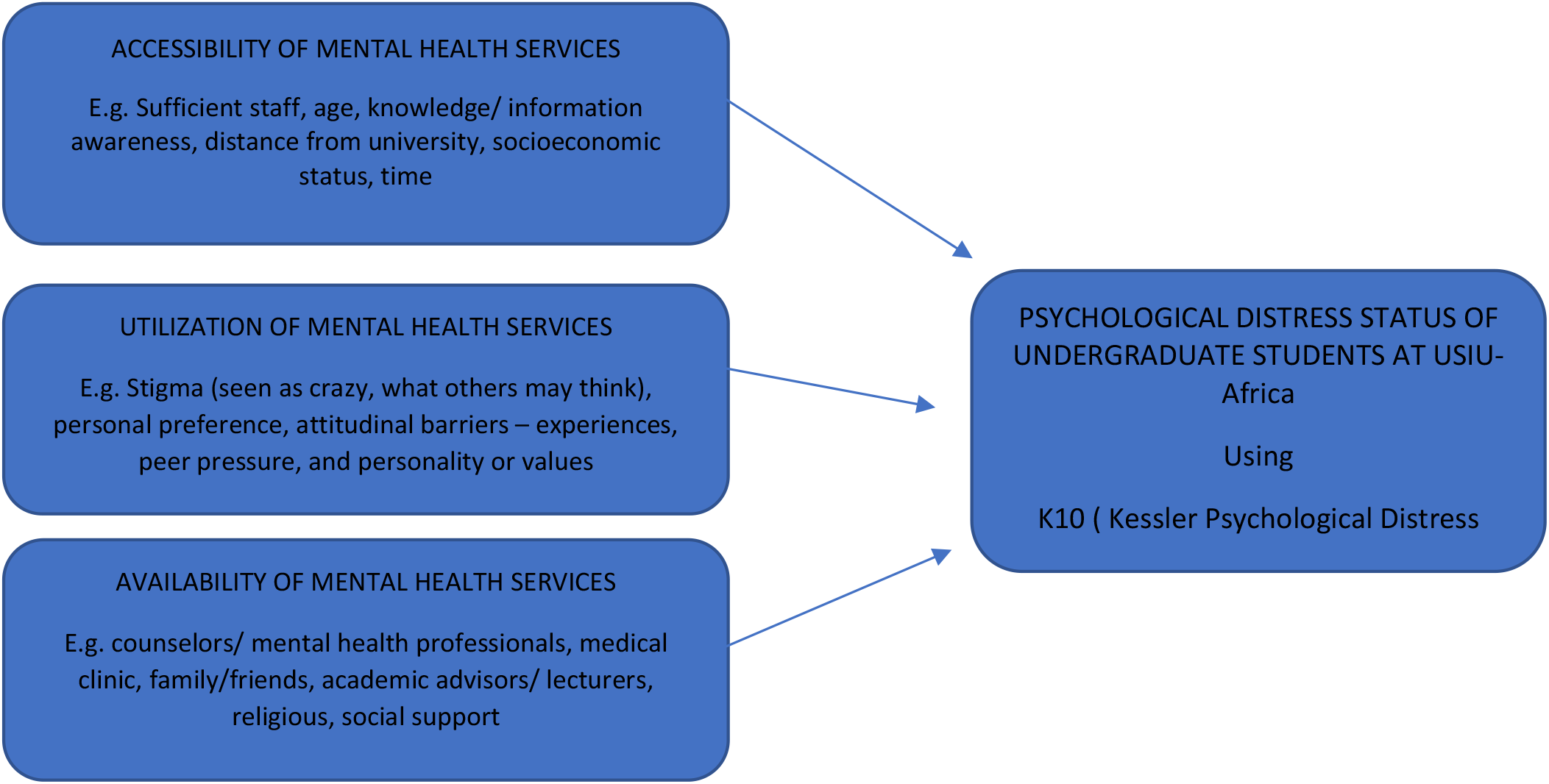

## 1.0 INTRODUCTON

Mental health is a significant component of a person’s well-being influencing how one operates psychologically, emotionally, and socially with oneself and others (WHO, 2022).Mental health plays a role in the physical health of a person. It affects one’s quality of life. Mental health condition is said to affect a quarter of people at some time in their lives (Friederich, 2017). Despite this fact, mental health remains to be a major public health concern globally. The prevalence has greatly been exacerbated by Covid-19 pandemic (Surapon, 2021). Mental disorders accounts for the highest burden of disease among young people worldwide, with depression being the highest disease burden (Kutcher et al., 2019). Kutcher and his colleagues reported that 70% of mental health disorders can be diagnosed before the age of 25 years which makes adolescent years crucial in providing youths with early and effective mental health services by the promotion of mental health.

University students are presented with great academic and social pressures that challenges their physical, social, and emotional areas of their lives while they explore the world outside the parental bubble. The prioritization of mental health of university students by promoting mental health services in learning institutions would increase positive social, emotional, psychological, thinking, and emotional strategies that would lay a foundation of better mental health and well-being for the future. While there has been some increase in research in the field of mental health of youths in Kenya, there is low level of availability, accessibility, and utilization of mental health services to university students. This may be due to inadequate prioritization of mental health by government, societal view of mental health as taboo, and lack of resources such as counsellors at higher education institutions.

Mental disorders do not discriminate against economic status, ethnic backgrounds, or gender. It is prevalent globally, affecting both developing and developed countries alike. Despite this fact, there remains a lacuna of mental health coverage in Africa as many resources are being channeled to tackle infectious diseases such as HIV/AIDS (Sankoh et al., 2018). There is limited availability and access to mental health services especially in low- and middle-income countries (Muhorakeye & Biracyaza, 2021). It is shocking that despite Kenyan’s youth (18-34 years) accounting for more than 60% of the country’s population, little to no strategies have been implemented to increase their access to mental health services. Research shows that the age group 18-29 years to be the highest proportion experiencing mental health issues as compared to the general population (Kessler et al., 2005). It is supported that 50% of mental health disorders develop by the age of 14 years and 75% by 24 years of age (Kessler et al., 2005). Thus, lack of action being taken to provide adequate mental health services to youths could have detrimental consequences in future.

Adolescents and young adults avoiding addressing their mental health issues could negatively impact them in their adulthood, affecting their health, education, family relations and economic situation (Sankoh et al., 2018). One study found that while adolescents in USA idealized suicide at 15%, developing countries in Sub-Saharan Africa have a higher risk of suicide idealization, with Kenya adolescent idealizing suicide at 27.9% (Were, 2020). This problem could be attributed to many factors such as lack of resources (e.g., counselors and funding) and societal view of mental health as a taboo. This major public health concern has resulted in a number of suicide cases among the youths, particularly university students. Without access to proper counseling and psychological help, university students struggle with poor academic performance, depression, and anxiety. Therefore, the purpose of this study was to provide evidence for development or enhancement of policies and strategies to promote mental health among the youth in institutions of higher learning. The main objective of the research was to determine the relationship between availability of mental health services, barriers to access to the services, and utilization of the services, and psychological distress status of undergraduate students at United States International University-Africa, Kenya (USIU-Africa). The specific objectives of the study include : (1) to determine psychological distress status of undergraduate students at United States International University-Africa, Kenya (USIU-A), (2) to determine available mental health services to undergraduate students at USIU-A and their influence on psychological distress status, (3) to determine barriers to accessibility of mental health services by undergraduate students at USIU-A and their association with psychological distress status, (4) to determine utilized mental health services by undergraduate students at USIU-A and their influence on psychological distress status.

Psychological distress refers to non-specific symptoms of stress, anxiety, and depression. Psychological distress can indicate impaired mental health and may reflect common mental disorders (Viertio, S et al, 2021). Research reports that psychological distress is connected to mental disorders. A study conducted by Edward and colleagues found that there is a dose relationship between extent of stress in each life area and increased odds of at least one mental disorder (Edward, A et al, 2010). Evidence found by research studies such as Canadian study in 16 universities found that the prevalence of psychological distress was higher among Ontario undergraduates(30%) compared to the general population. Additionally, the study reported that psychological distress varied by sex, gender, academic orientation was significantly related to elevated distress (Adalf, E et al;2001). A similar study found that 21.6% of undergraduates suffered from psychological distress and factors such as year of study were associated with mental distress. A second-year student was less likely ot suffer from psychological distress (Dessie,Y et al, 2013).

## 2.0 METHODS

This study was conducted between July 1, 2022, and August 2, 2022. Institutional Research Ethics Review Committee approval was sought from the United States International University – Africa (USIU-A/IRB/322-2022) and ethical clearance from the National Commission for Science, Technology, and Innovation (NACOSTI) was received before the start of data collection.

### 2.1 SETTINGS AND SUBJECTS

Participants in this study were recruited from United States International University – Africa which is a private university located in Nairobi, Kenya. The target population is youths in institutions of higher learning. The study population is aisundergraduate students who are actively enrolled during Summer 2022 semester at United States International University-Africa, Kenya.

### 2.2 DATA COLLECTION

This study employed a mixed methods research approach, specifically using concurrent qualitative and quantitative approaches. Data were collected from a survey of undergraduate students’ perceptions of mental health services offered at the university and validated by interviews with counselors at USIU-A. An exploratory sequential mixed method research design was used. In the quantitative arm, we used cross-sectional survey questionnaires to assess the psychological distress status of the undergraduatee students. The questionnaire was also used to assess students’ perception of the availability, accessibility, and utilization of mental health services at the university. In the qualitative arm, key informants were interviewed by counselors to validate the survey data.

Both cluster sampling and Stratified sampling were used to select the sample from the population. cluster sampling was used in the qualitative component and stratified sampling was used in the quantitative component of the study. After dividing the population into strata for each school in the university. The five academic schools as the of summer semester 2022 included: School of Business (1252), School of Humanities and Social Sciences (956), School of Pharmacy and Health Sciences (404), School of Science and Technology (1002), and School of Communications, Cinematics, and Creative Arts (350). The total number of undergraduate students in USIU-A was determined as 3964 undergraduate students. Selecting students in each school, it allowed for the topic under study to be compared for different subgroups of the population, findings being more meaningful.

Administration records were obtained from the Registrar at USIU-A to determine a list of classes being taught during the semester. Classes from each school were randomly selected to be included in the study. Google Forms was used to create the survey questionnaire. A physical copy of the surveys was shared in person with students in the randomly selected classes in the schools at USIU-Africa between July 1, 2022, and August 2, 2022. Those who preferred to fill survey virtually and had consented to participate were sent a link to the survey via WhatsApp and Gmail. Data collection began by engaging in a discussion with a mental health professional at the university to determine the types of mental health services available at the University. After determination of the study sample size, the questionnaire was designed to capture the three main components of mental health services: availability, accessibility, and utilization. To enhance the response rate, lecturers were asked to set aside a small portion of their class time to allow for the selected class to participate in the study.

### 2.3 VARIABLES

This research looked at the relationship between student’s psychological distress status (one independent variable) and three categories of dependent variables: knowledge of the availability of mental health services, whether available MHSs are accessed or not, and whether accessible MHSs are utilized or not by undergraduate students at the United States International University – Africa, Kenya. The American Psychological Association (APA) defines mental health services as “any intervention-assessment, diagnosis, treatment, or counseling offered in private, public, inpatient, or outpatient settings for the maintenance or enhancement of mental health or the treatment of mental or behavioral disorders in individual and group contexts (APA, 2022). The three dimensions of MHS examined in this study are operationally defined as follows. Availability of mental health service refers to the existence of the service whether it is accessed or not. Mental health service may be available at a facility (e.g., hospital, health center, or clinic) on- or off-campus or it may be in the form of mental health information. Accessibility of mental health services refers to the extent to which students seek the service regardless of whether it is received or not. Activation of mental health service refers to the actual use of the service, for example, receiving psychological counseling therapy.

### 2.3 ANALYSES AND TOOLS

K10 (Kessler Psychological Distress Scale), a 10-item questionnaire intended to yield a global measure of distress based on anxiety and depression symptoms experienced was adapted for use in this study. Studies have shown K10 to be of high validity and reliability to measure psychological distress (Ongeri, L et al; 2022). K10 was used in a study conducted by Inanova and colleagues to measure psychological distress status by screening for psychological distress. They scored individuals with scores ≥30 to be suffering from severe psychological distress (Ivanova, O et al; 2022). In the present study, scores were interpreted as follows: a score under 3 is more likely to suffer from psychological distress and a score of greater than or equal to 30 to likely to not suffer from psychological distress. SPSS Statistics 28.0 was used to analyze data and logistic regression determine was used to examine the relationship between student’s psychological distress status (one independent variable) and three categories of dependent variables: knowledge of the availability of mental health services, whether available MHS are accessed or not, and whether accessible MHS are utilized or not by undergraduate students at United States International University – Africa, Kenya. Findings were summarized using Excel graphs and charts.

## 3.0 RESULTS

### 3.1 SAMPLE POPULATION

Participants surveyed included more females (57%) than males, the majority belonged to the 21-25 years age group (57.4%), resided far from campus (46.6%), were in their senior year (33.7%), and belonged to the School of Business (32.9%) (Table 1).

**Table 1.** Participant characteristics

| Variables |  | Number (n) | Percentage (%) |
| --- | --- | --- | --- |
| Gender |  |  |  |
| Female |  | 142 | 57.00% |
| Male |  | 107 | 43.00% |
| Age group |  |  |  |
| 21-25 years |  | 143 | 57.40% |
| 15-20 years |  | 65 | 26.10% |
| 26-35 years |  | 38 | 15.30% |
| 36-45 years |  | 3 | 1.20% |
| Residence |  |  |  |
| Far from campus |  | 116 | 46.60% |
| Near campus |  | 115 | 46.20% |
| On campus |  | 18 | 7.20% |
| Year of study |  |  |  |
| Senior |  | 84 | 33.70% |
| Freshman |  | 70 | 28.10% |
| Sophomore |  | 40 | 16.10% |
| Junior |  | 41 | 16.50% |
| Senior (5<sup>th</sup> year) |  | 12 | 4.80% |
| School |  |  |  |
| School of Business |  | 82 | 32.90% |
| School of Humanities and Social Science |  | 54 | 21.70% |
| School of Pharmacy and Health Sciences |  | 51 | 20.50% |
| School of Science and Technology |  | 49 | 19.70% |
| School of Communications, Cinematics, and Creative Arts |  | 13 | 5.20% |

### 3.2 PSYCHOLOGICAL DISTRESS STATUS OF RESPONDENTS

Using the K10 psychological distress scale, about three-quarters (76.8%) of the respondents suffered from psychological distress, and about one-fifth (21.3%) were not to be suffering (Table 2).

**Table 2.**
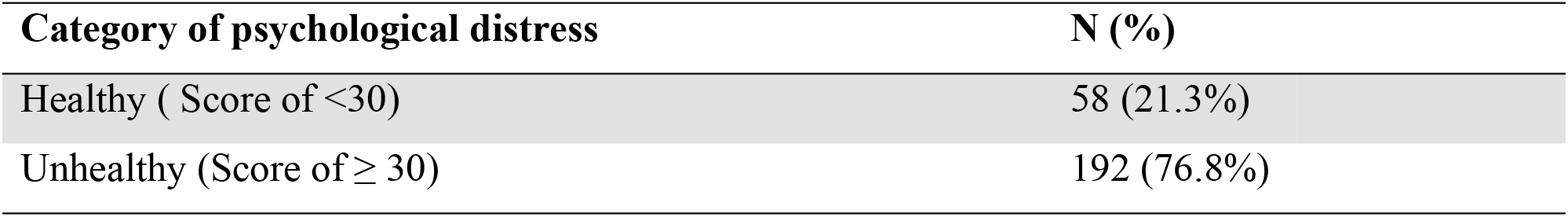
Psychological distress status of participants

| Category of psychological distress | N (%) |
| --- | --- |
| Healthy ( Score of <30) | 58 (21.3%) |
| Unhealthy (Score of $\geq 30$) | 192 (76.8%) |

The chi-square test showed that there were no statistically significant differences in psychological distress status by residence (*p* = 0.301), age group (*p* = 0.073), or year of study (*p* = 0.107). However, psychological distress status varied by gender(χ^2^ (df=2) = 13.452, *p* = 0.01)and school (χ^2^ (df=2) =11.379, *p* = 0.044). For example, female students were more likely to be suffering from psychological distress compared to male students (Table 3).

**Table 3.**
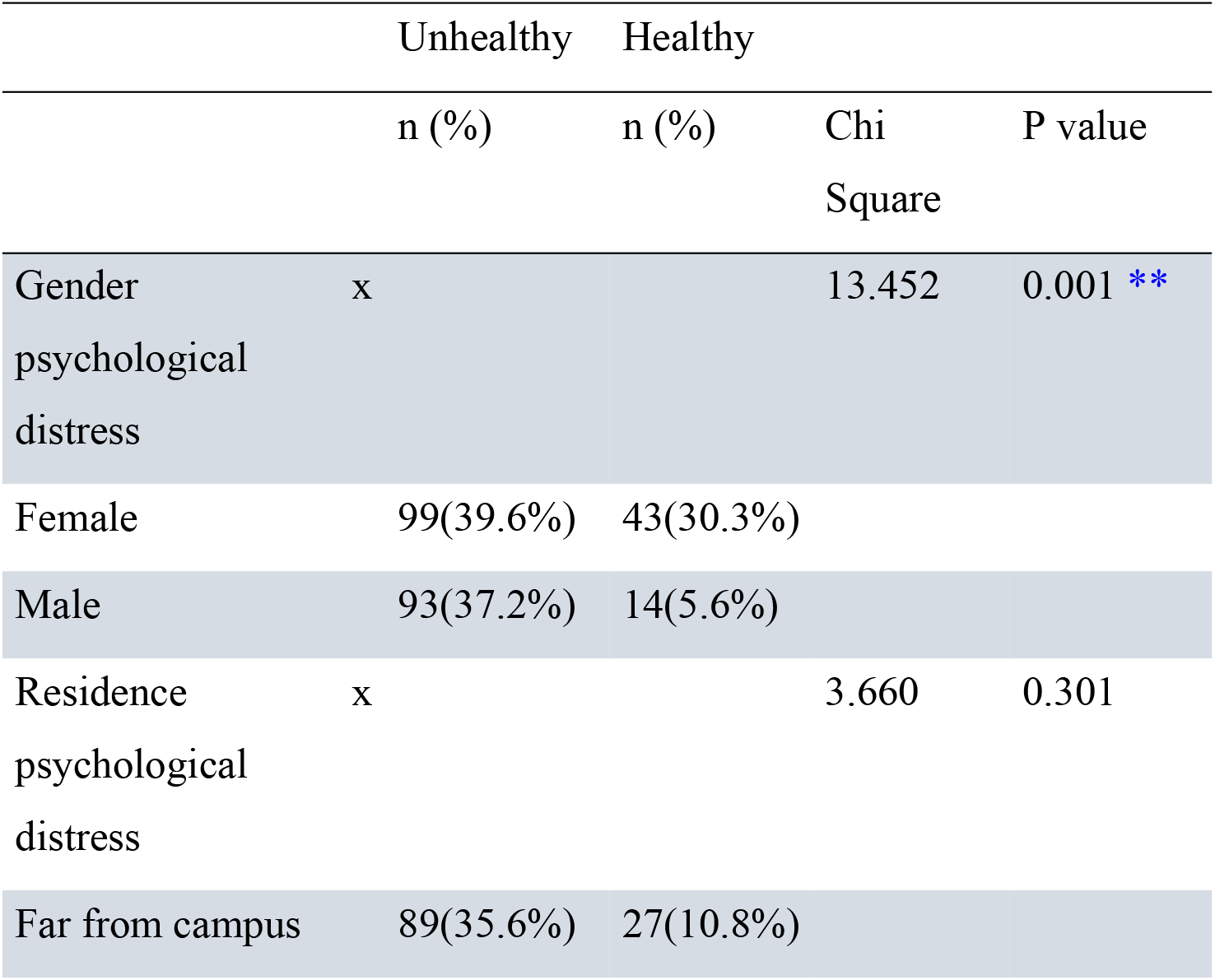
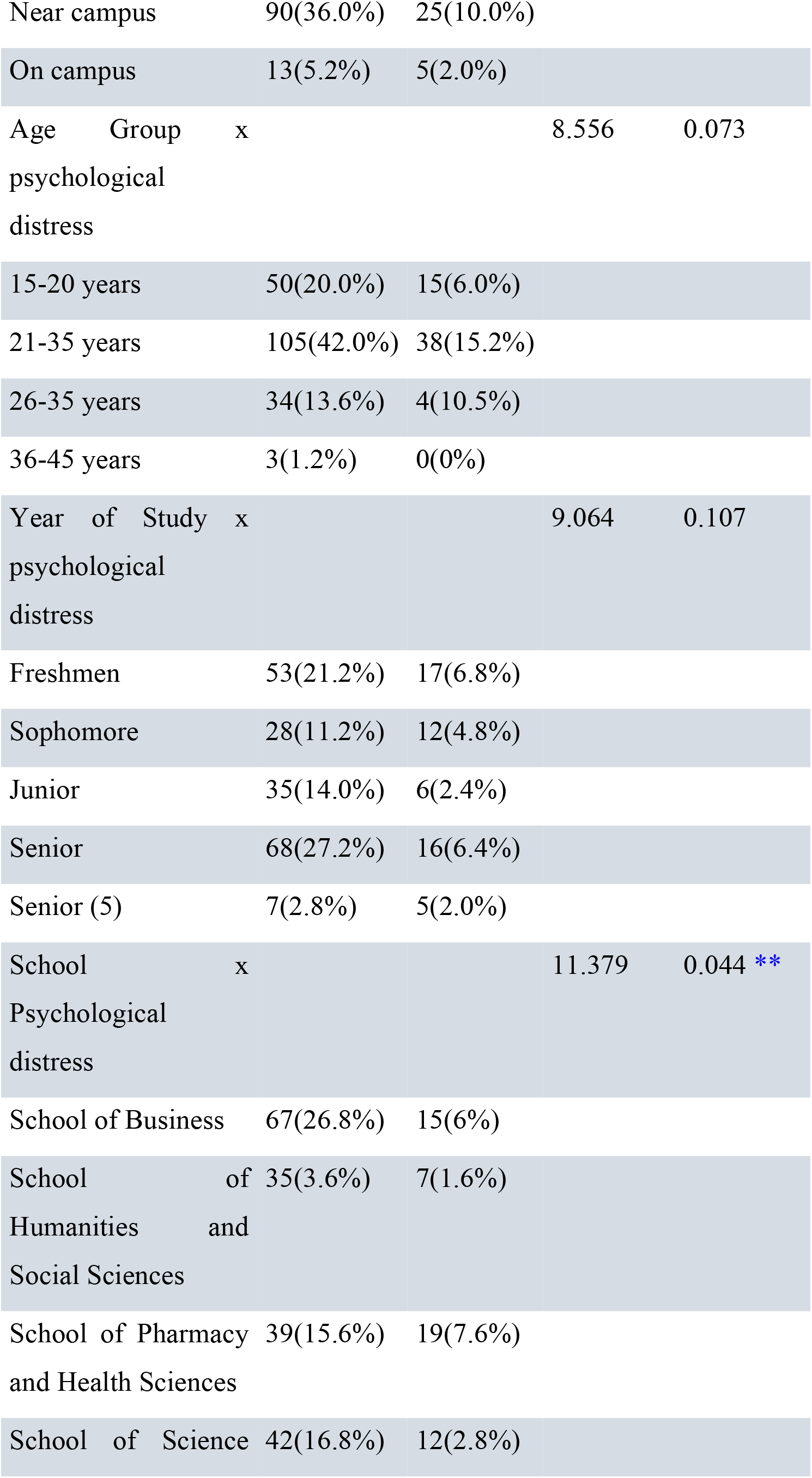
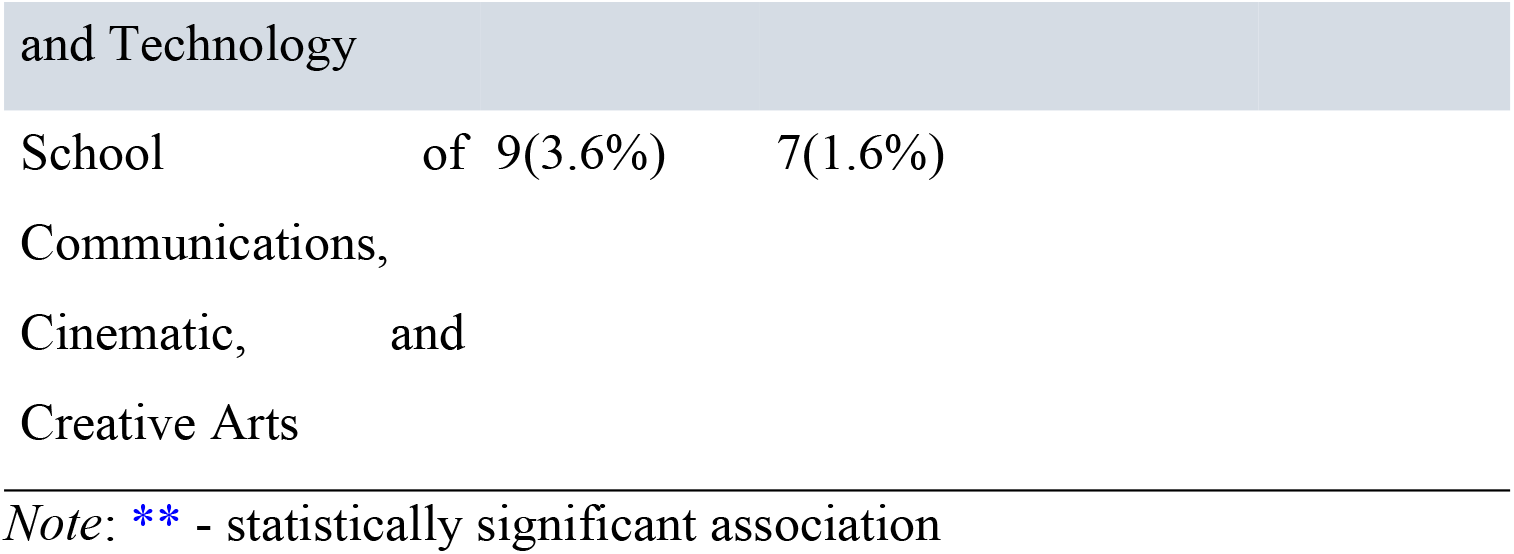
Association between participant characteristics and psychological distress status

## AVAILABILITY, ACCESSIBILITY, AND UTILIZATION OF MENTAL HEALTH SERVICES

### 3.4 AVAILABLE MENTAL HEALTH SERVICES

Based on interviews with four counselors at the Counseling Center at USIU-Africa, the following seven sources of mental health services are available at USIU-A: 1) counselors, 2) medical clinic, 3) social support such as friends and relatives, 4) lecturer/course advisors, 5) peer counselors, 6) personal coping skills, and 7) psychologists. However, they stated that religious leaders, psychiatrists, pharmacies, and use of medication were not available for students as sources of mental health services.

With respect to students surveyed, the majority indicated being aware of the following sources of mental health services as available at USIU-A: counselor (86.7%), social support (83.5%), peer counselor (80.3%), medical clinic (79.9%), and lecturer/course advisor (79.1%) (Table 4). Interestingly, fewer students seemed to be aware of psychologists as an available source of mental health service, let alone personal coping strategies, 67.9%, and 59.4% respectively.

**Table 4.**
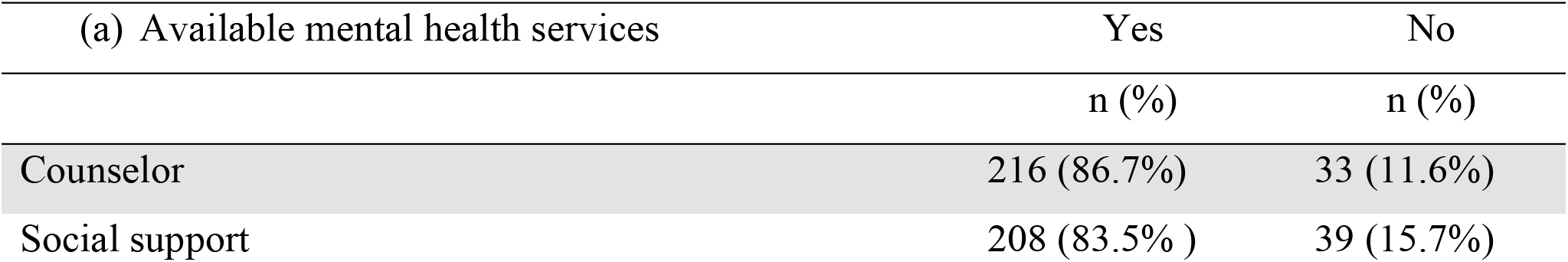
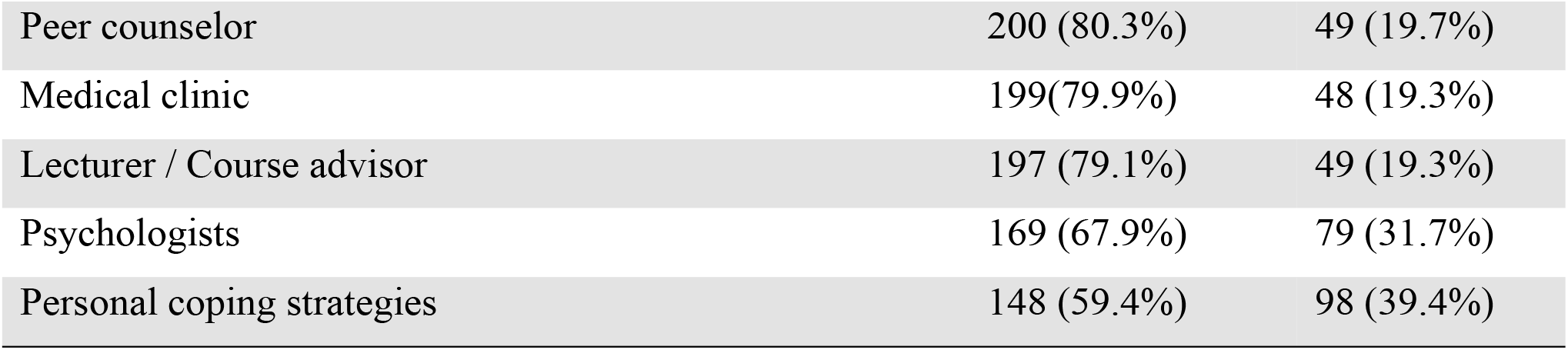
Available mental health services to USIU-A students

| (a) Available mental health services | Yes | No |
| --- | --- | --- |
|  | n (%) | n (%) |
| Counselor | 216 (86.7%) | 33 (11.6%) |
| Social support | 208 (83.5%) | 39 (15.7%) |
| Peer counselor | 200 (80.3%) | 49 (19.7%) |
| Medical clinic | 199(79.9%) | 48 (19.3%) |
| Lecturer / Course advisor | 197 (79.1%) | 49 (19.3%) |
| Psychologists | 169 (67.9%) | 79 (31.7%) |
| Personal coping strategies | 148 (59.4%) | 98 (39.4%) |

### 3.5 INFLUENCE OF MENTAL HEALTH SERVICES AVAILABILITY ON STUDENTS’ PSYCHOLOGICAL DISTRESS STATUS

No statistically significant association was found between psychological distress status and students’ awareness of the availability of the following mental health services: social support (*p* = 0.097), counselor (*p* = 0.100), medical clinic (*p* = 0.164), lecturer/course advisor (*p* = 0.180), and peer counselor (*p* = 0.053) (Table 5). However, psychological distress status varied by students’ awarenessthe of availability of psychologists (χ^2^ (df=2)= value, *p* = 0.005) or personal coping strategies (*p* = 0.018).

**Table 5.** Influence of availability of mental health services on psychological distress status

| Available Services | Unhealthy | Healthy | Chi square | P value |
| --- | --- | --- | --- | --- |
| Psychologist | 122(48.8%) | 47(18.8%) | 10.467 | 0.005** |
| Personal coping strategies | 107(42.8%) | 41(16.4%) | 8.064 | 0.018 ** |
| Social support | 164(65.6%) | 45(18.0%) | 4.675 | 0.097 |
| Counselor | 164(65.6%) | 52(20.8%) | 4.603 | 0.100 |
| Medical clinic | 152(60.8%) | 47(18.8%) | 3.617 | 0.164 |
| Lecturer / Course advisor | 151(60.4%) | 46(18.4%) | 3.435 | 0.180 |
| Peer counselor | 150(60.0%) | 50(20.0%) | 5.859 | 0.053 |

Note: \*\* - statistically significant association

### 3.6 BARRIERS TO ACCESSIBILITY OF MENTAL HEALTH SERVICES

Amongst the respondents surveyed, on average, the majority cited *attitudinal barriers* as preventing them from accessing mental health services. This was followed by *stigma barriers* (31.0%) and *instrumental barriers* (27.6%). The most frequently cited barriers in each of the three categories were as follows: attitudinal: “self-sufficiency (86.7%)” and “Thinking the problem will get better” (80.3%); stigma-related: “peers (40.6%)” and “society (40.6%)”; and instrumental: “time (38.6%)” and “disbelief in mental health (28.5%).”

Based on interviews with four counselors, it emerged that the main barriers to seeking mental health services at USIU-A are *stigma* (e.g., self-stigma, stigma from peers, and stigma from society) and *mental health literacy*. Emphasizing the impact of stigma, one counselor reported, “*The topic of mental health has gained notoriety over the last two years, yet stigma remains a key limitation. Students possess the fear of being labeled. Being seen walking into the Counseling Center is dreaded*.” Another counselor noted, “*Students may feel that they may be stigmatized by society and seen as being crazy*.” Two counselors identified a lack of knowledge about mental health issues as a barrier, stating “*Some* [students] *don’t know the symptoms and don’t know their* [mental health] *issues or minimize their issues*.” In addition, cultural limitations were stated as preventing some students from accessing mental health services.

### 3.7 ASSOCIATION BETWEEN BARRIERS TO ACCESSIBILITY TO MENTAL HEALTH SERVICES AND STUDENTS’ PSYCHOLOGICAL DISTRESS STATUS

Cramer’s V Analyses of survey data showed that there were no statistically significant associations between barriers to access to mental health services and students’ psychological distress status: self-stigma(p=0. 805), cultural stigma(p=0. 471), gender of a counselor (p=0. 258), confidentiality(p=0.476), Attitudinal barriers: don’t like to talk emotions (p=0.833), health system mistrust (p=0.548), thought the problem would get better(0.668), negative past experiences (0.801), prefer alternative care (p=867) and instrumental barriers: mental health literacy (0.164), time(0.540), location(0.288), and disbelief in mental health (0.155) (Table 7). In other words, students who cited the above stigma, attitudinal, or instrumental barriers did not differ in their psychological distress status.

**Table 6.**
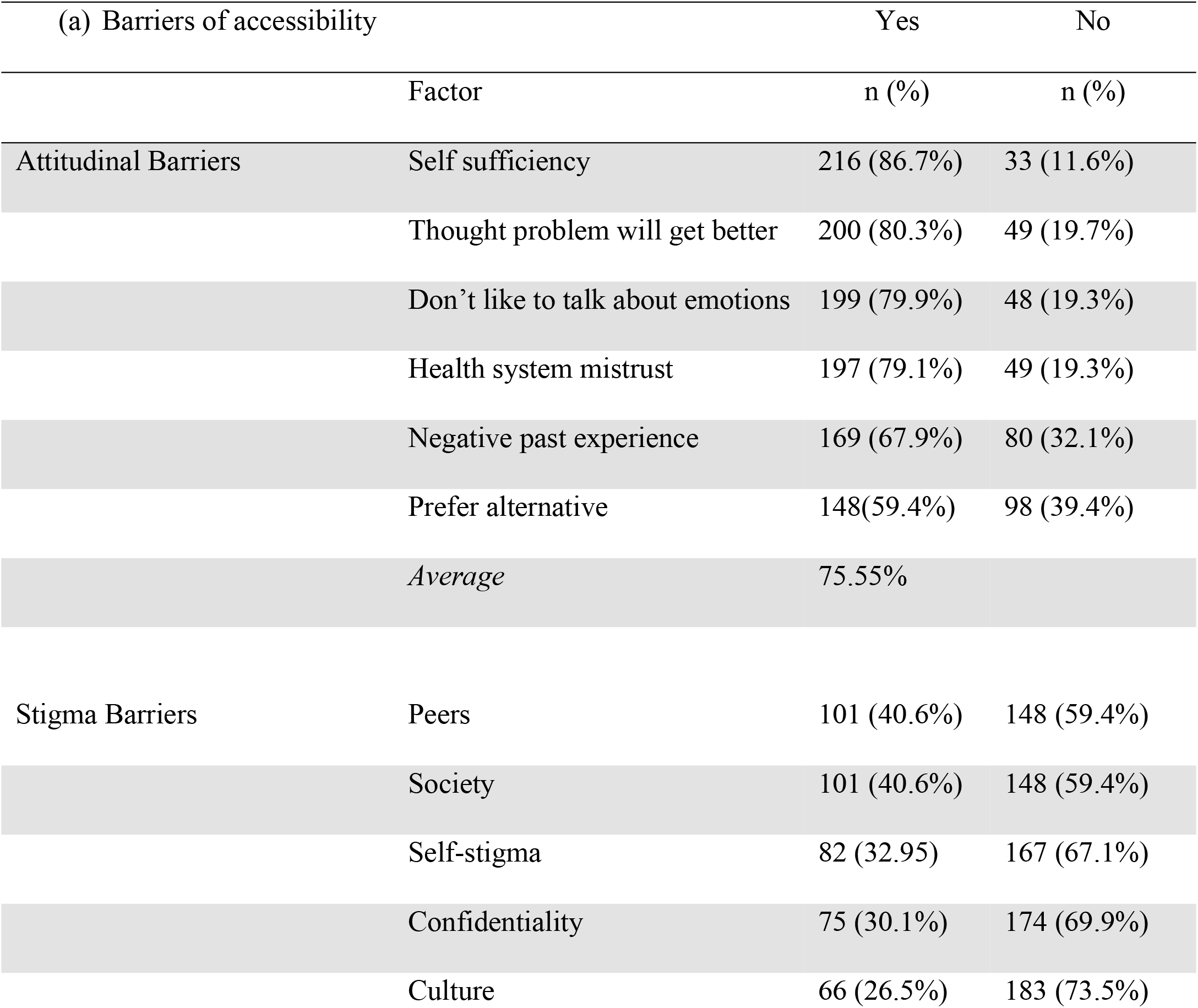
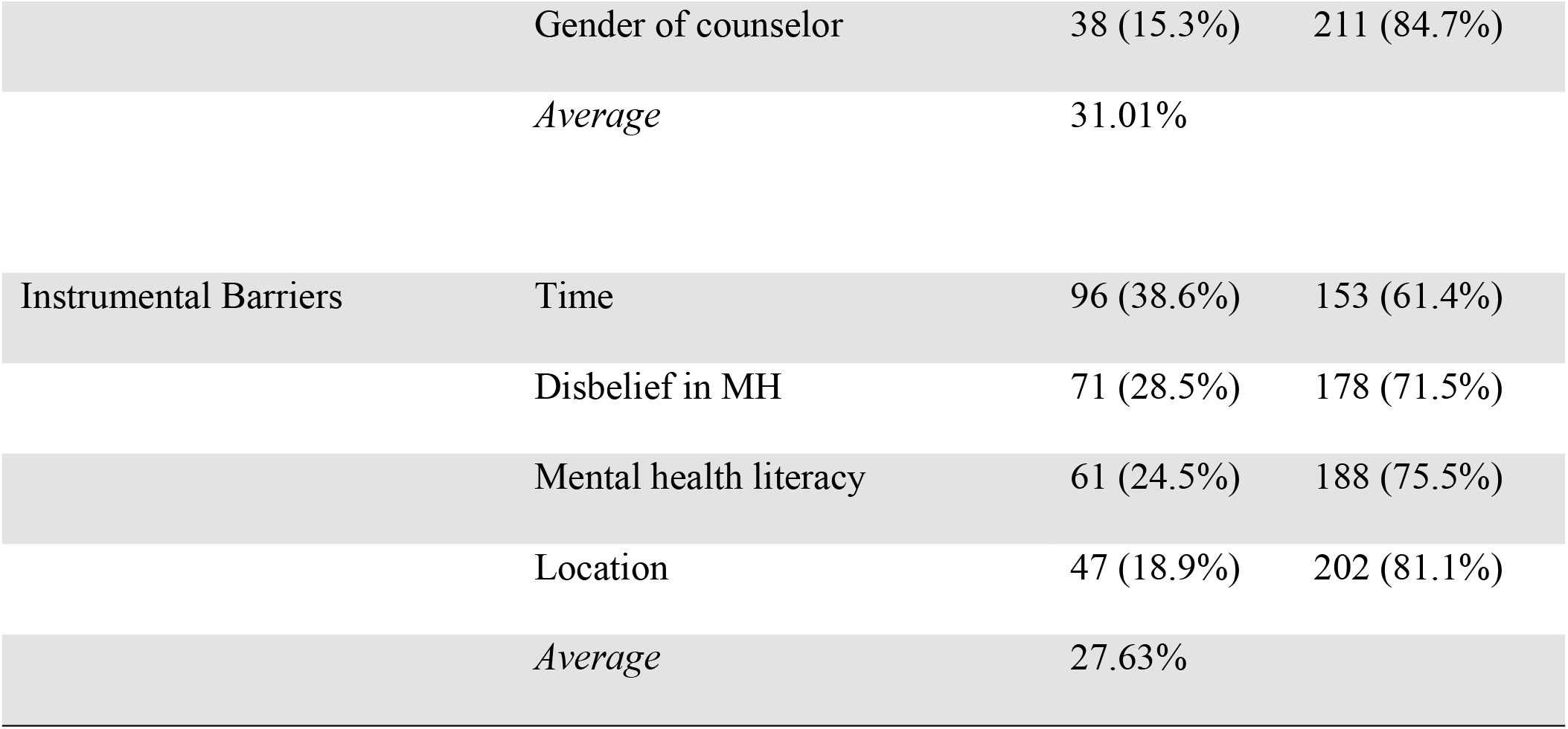
Barriers to access to mental health services by USIU-A students

**Table 7.** Association between barriers to access to mental health services and psychological distress status

| Barriers | Unhealthy<br>n (%) | Healthy<br>n (%) | Cramer's v | (p value) |
| --- | --- | --- | --- | --- |
| Stigma barriers |  |  |  |  |
| Peers | 87(34.9%) | 14(5.6%) | 0.178 | 0.05 ** |
| Society | 87(34.9%) | 14(5.6%) | 0.178 | 0.005 ** |
| Self-stigma | 64(25.7%) | 18(7.2%) | 0.016 | 0.805 |
| Culture | 53(21.3%) | 13(5.2%) | 0.046 | 0.471 |
| Gender of counselor | 32(12.9%) | 6(2.4%) | 0.072 | 0.258 |
| Confidentiality | 60(24.1%) | 15(6.0%) | 0.045 | 0.476 |
| Attitudinal barriers |  |  |  |  |
| Self sufficiency | 117(47.0%) | 26(10.4%) | 0.130 | 0.040 ** |
| Don't like to talk emotions | 98(39.4%) | 30(12.0%) | 0.013 | 0.833 |
| Health system mistrust | 89(35.7%) | 29(11.6%) | 0.038 | 0.548 |
| Thought problem will get better | 78(31.3%) | 25(10.0%) | 0.028 | 0.663 |
| Negative past experience | 40(16.1%) | 11(4.4%) | 0.016 | 0.801 |
| Prefer alternative care | 39(15.7%) | 11(4.4%) | 0.011 | 0.867 |
| Instrumental barriers |  |  |  |  |
| Mental health literacy | 51(20.5%) | 10(4.0%) | 0.088 | 0.164 |
| Time | 76(30.5%) | 20(8.0%) | 0.039 | 0.540 |
| Location | 39(15.7%) | 8(3.2%) | 0.067 | 0.288 |
| Disbelief in MH | 59(23.7%) | 12(4.8%) | 0.090 | 0.155 |
| Too unwell to seek MH | 15(6.0%) | 6(2.4%) | 0.041 | 0.517 |

Note: \*\* - statistically significant association

However, there were statistically significant associations between barriers to access to mental health services and students’ psychological distress status: peer stigma (v=0.178, p=0.05), societal stigma (v=0.178, p=0.005), and self-sufficiency (v=0.130, 0.040).

### 3.8 UTILIZED MENTAL HEALTH SERVICES

Based on analysis of survey data, the top three mental health services that were identified as being utilized by students were: *lecturer/course advisor* (79.1%), *psychologists* (67.9%), and *social support* (48.2%) (Table 8). The least cited source of mental health service being utilized by students was peer counselors (21.7%).

**Table 8.** Utilization of mental health services by USIU-A students

| (b) Utilized mental health services | Yes | No |
| --- | --- | --- |
|  | n (%) | n (%) |
| Lecturer / Course advisor | 197 (79.1%) | 49 (19.3%) |
| Psychologist | 169 (67.9%) | 80 (32.1%) |
| Social support | 120 (48.2% ) | 129 (51.8%) |
| Personal coping strategies | 105 (42.2 %) | 144 (57.8%) |
| Counselor | 86(34.5%) | 163(65.5%) |
| Medical clinic | 79(31.7 %) | 170 (68.3%) |
| Lecturer/Course Advisor | 70(28.1%) | 179(71.9%) |
| Peer counselor | 54 (21.7%) | 195 (78.3%) |

Based on interviews with counselors, it emerged that there was low utilization of counseling services at the university. One counselor stated that *“It is recommended that 10% of a population using services available is internationally accepted* [by WHO]. *I would not say a specific percentage, but I can say that less than 10% of undergraduates use counseling services*.”

### 3.9 INFLUENCE OF MENTAL HEALTH SERVICES UTILIZATION ON STUDENTS’ PSYCHOLOGICAL DISTRESS STATUS

Cramer’s v analyses of survey data revealed that there were no statistically significant differences in psychological distress status by students’ utilization of mental health services: personal coping strategies (p = 0.768), social support (p = 0.445), counselors (p = 0.921), medical clinic (p = 0.725), lecturer/course advisor (p = 0.310), and peer counselor (p = 0.618) (Table 9).

**Table 9.**
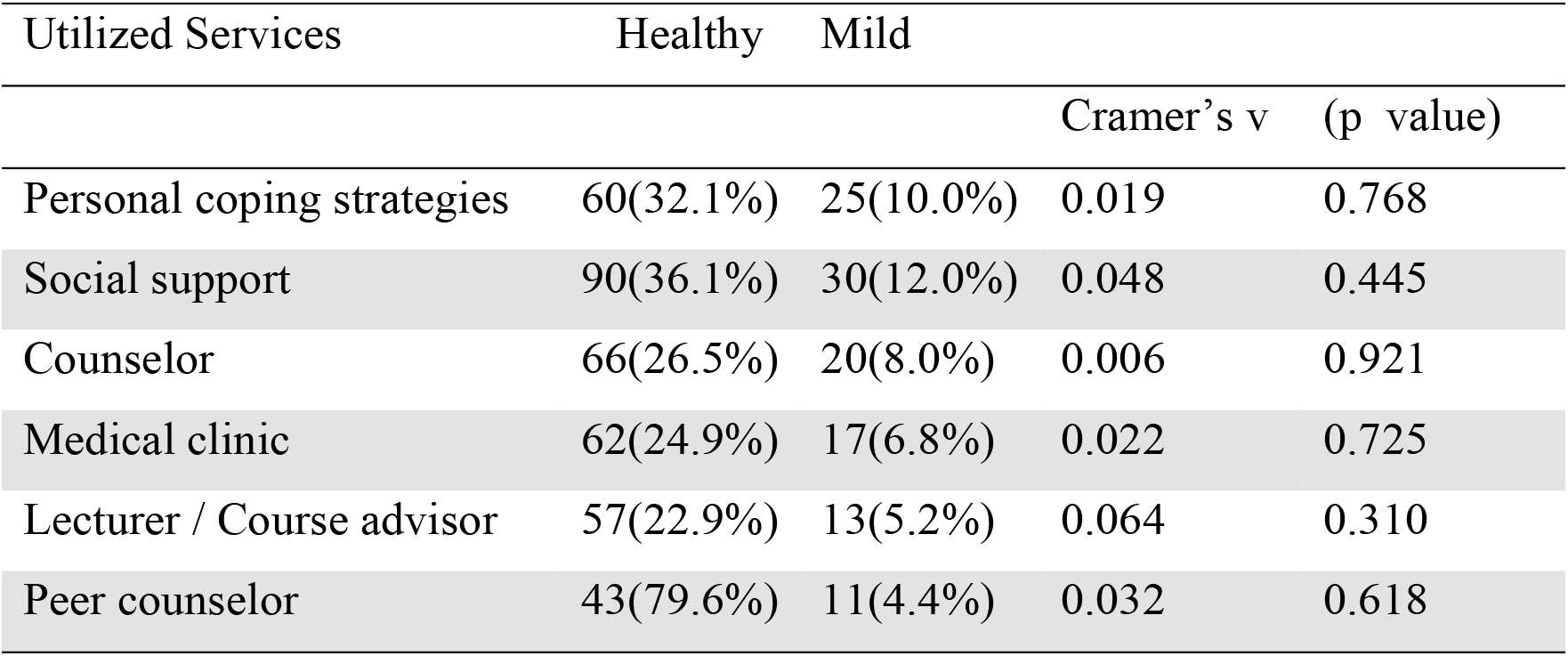
Influence of utilization of mental health services on psychological distress status of USIU-A students.

## 4.0 DISCUSSION AND CONCLUSION

This study has found meaningful findings that will be useful in improving the psychological distress status of undergraduate students focusing on mental health services. It was found that majority of USIU-Africa undergraduates students suffer from psychological distress. This is much greater than the findings from research conducted by Dessie and colleagues (Dessie, Y et al., 2013) which reported that 21.6% of undergraduate students suffered from psychological distress The two studies differed in several factors-: the study population, assessment tool used to assess psychological distress, and sampling procedure. Nonetheless, this study emphasizes that psychological distress prevalent among university students in Africa.

There were statistically significant differences in students’ psychological distress status and characteristics such as gender and school. Several sources of mental health services (counselors, medical clinic, social support, lecturer/course advisor, peer counselor, personal coping strategies, and psychologists) are available at this institution as identified from the key informant interview with counselors. Majority of the students were aware of these services available with majority of students aware of counselor’s availability. There was a statistically significant difference noted between students’ awareness of psychologists or personal coping strategies and students’ psychological distress status.

The most frequently cited barrier preventing students’ from accessing mental health services at the institution was attitudinal barriers with the highest categories being feelings of self-sufficiency and “thinking the problem will get better”. Statistical association was found between students’ psychological distress status and barriers to accessibility of mental health services such as peer stigma, societal stigma, and self-sufficiency. It was reported that students’ top three most utilized mental health services were lecturers/ course advisors, psychologists, and social support. From the interview conducted, it was apparent that there was a low utilization of mental health services by students. No statistically significant difference was found between psychological distress status and students’ utilization of mental health services. The major findings from this study informs us of key components of mental health services that must be considered when improving the mental health services available. It was reported that mental health services are available to students and students are aware of them. Although majority of students were aware of counselors as mental health service available at the university, the most utilized mental health service were lecturers/course advisors, psychologists, and social support. This study corresponds to other studies in that it concludes that there is low level of mental health utilization by college students. Attitudinal barriers were the most frequently cited as preventing students from accessing mental health services.

A few limitations of this study that must be considered are findings of this study are based on a relatively small sample size, 249 students out of expected 351 students. Perhaps, with a larger sample size, some of the findings may be different.

This study recommends future research to focus on not only in determination of the association between the three components of mental health services (availability, accessibility, and utilization) and psychological distress status of college students but also the strength of association. Additionally, rather than using a cross-sectional design, a longitudinal study design can be employed to determine how psychological distress status change through academic years. This can help the university decide not only the best to time to intervene but also the most effective mental health service to provide. The findings from this study suggest that there is need to broaden or enrich available mental health services by offering alternative forms of support to college students. These include, for example, using social support groups in therapeutic activities such as painting or music and strengthening personal copying skills by having programs in university on stress management and tools available. The policy should be implemented to facilitate student knowledge of mental health disorders and the importance of mental health services through social media to reduce the level of societal stigma.

## Data Availability

Data will be available upon request.

## ACKNOWLEDGEMENTS

I would like to acknowledge the support of the United States International University – Africa (USIU-Africa), for funding, USIU – Africa counseling center, undergraduate students, and counselors involved in the study.

